# Deep Neural Networks for Human’s Fall-risk Prediction using Force-Plate Time Series Signal

**DOI:** 10.1101/2021.05.19.21257466

**Authors:** M. Savadkoohi, T. Oladunni, L.A. Thompson

## Abstract

Early and accurate identification of the balance deficits could reduce falls, in particular for older adults, a prone population. Our work investigates deep neural networks’ capacity to identify human balance patterns towards predicting fall-risk. Human balance ability can be characterized based on commonly-used balance metrics, such as those derived from the force-plate time series. We hypothesized that low, moderate, and high risk of falling can be characterized based on balance metrics, derived from the force-plate time series, in conjunction with deep learning algorithms. Further, we predicted that our proposed One-One-One Deep Neural Networks algorithm provides a considerable increase in performance compared to other algorithms. Here, an open source force-plate dataset, which quantified human balance from a wide demographic of human participants (163 females and males aged 18-86) for varied standing conditions (eyes-open firm surface, eyes-closed firm surface, eyes-open foam surface, eyes-closed foam surface) was used. Classification was based on one of the several indicators of fall-risk tied to the fear of falling: the clinically-used Falls Efficacy Scale (FES) assessment. For human fall-risk prediction, the deep learning architecture implemented comprised of: Recurrent Neural Network (RNN), Long-Short Time Memory (LSTM), One Dimensional Convolutional Neural Network (1D-CNN), and a proposed One-One-One Deep Neural Network. Results showed that our One-One-One Deep Neural Networks algorithm outperformed the other aforementioned algorithms and state-of-the-art models on the same dataset. With an accuracy, precision, and sensitivity of 99.9%, 100%, 100%, respectively at the 12th epoch, we found that our proposed One-One-One Deep Neural Network model is the most efficient neural network in predicting human’s fall-risk (based on the FES measure) using the force-plate time series signal. This is a novel methodology for an accurate prediction of human risk of fall.

## I. Introduction

The detection of fall-risk linked balance impairment is of significant societal relevance. Falling has been reported as the second most common reason of accidental injuries and death, with road traffic accidents being the first (World Health Organization-Falls, n.d.). Treating fall-related injuries is extremely costly. In the United States, costs related to fatal and nonfatal falls in year 2000 was estimated as $0.2 billion and $19 billion respectively for the adults over the age of 65 (Stevens et al., 2006). This number increased to $50 billion in 2015 (Florence et al., 2018). Balance impairment can have devastating effects on all individuals; though, particularly, older individuals are a vulnerable population. Thus, studying the balance characteristics and comparing the underlying patterns of balance attributes could be significantly helpful in predicting fall and reduce its associated risk.

Balance aids us in maintaining stability and preventing falls during a variety of daily activities that we often take for granted, such as getting up out of bed or from a chair, standing, and walking (Balance Disorders — Causes, Types & Treatment | NIDCD, n.d.; Winter, 1995). However, musculoskeletal, vestibular, visual, somatosensory, and proprioceptive disorders can lead to balance disorders in wide range of people (Balance Problems - Symptoms and Causes - Mayo Clinic, n.d.; Pialasse et al., 2016). Numerous studies have shown that older people, in particular, are more affected by balance impairment (Abrahamová D, 2008; Alexander, 1996; Brooke, 2010). Older adults are more prone to balance impairment due to the natural process of aging which results in degeneration of the above systems, confounded by muscle weakness and other age-related disorders, such as Parkinson’s disease, Stroke, Chronic Tremor, Multiple Sclerosis, and osteoporosis. Older people who suffer from these impairments are highly prone to fall and fall-related injuries (Alshammari et al., 2018; Beghi et al., 2018; Ozcan et al., 2005; Thompson et al., 2018).

Aside from physical injury due to falling, according to the National Institute of Aging (NIA), fear of falling increases as people age. As a result, older people tend to isolate themselves and avoid regular social activities (Prevent Falls and Fractures | National Institute on Aging, n.d.). This situation leads to other physical and mental disorders (e.g., decrease in bone mineral density, fractures, loss of independence, loss of confidence, anxiety, depression, panic attacks). Studies have shown that 13% of adults experience imbalance during the age range of 65-69 years old and this number increases to 46% percent after the age of 85 (Burns & Kakara, 2018; Osoba et al., 2019).

This paper is organized as follows: Section II overviews the available literature and highlights some of the studies conducted on the human balance and gait characteristics; Section III explains the materials and methods for the study and our methodology; Section IV explains the experimental results and V our discussions. We present our conclusions in Section VI.

## II. Literature Review

Researchers have employed various techniques to record human static and dynamic balance characteristics towards the ultimate goal of preventing falls. Using various equipment and analytical methods, scientists have gained a broad perspective towards human balance underlying patterns and factors that might affect the ability of humans in having a safe and normal life.

Kinematics and kinetics metrics are valuable indicators of human balance and postural control and are acquired using technologies, such as motion capture, force-plate, electromyography (EMG), sensors, accelerometers (Winter, 1995). The force-plate is perhaps the most popular equipment used to measure standing balance and gait. To identify and treat balance disorders, appropriate identification methods must be employed. Physicians and/or physical therapists often use conventional clinical methods to determine what is considered “good” or “poor” balance.

Clinical tests evaluate postural control of people and their stability (balance) while doing several static and/or dynamic motor tasks. Some examples are Activities-Specific Balance Confidence (ABC), Berg Balance Scale (BBS), Timed Up to Go test (TUG), Fall Efficacy Scale (FES) and Balance Error Scoring System (BESS). The advantages of these assessments are that they are easy to use, require minimal (if any) equipment, are quick to administer and inexpensive. However, they provide limited information to quantify the underlying balance metrics and provide a general understanding of balance ability.

Force-plates are practical devices used in research and clinical studies to characterizes gait, balance, and other biomechanics features. Sport and performance monitoring, balance impairment detection, occupational safety and motion analysis are some of the main applications of this measuring instrument. Time series signals recorded by force-plate sensors from which features, such as Center of Pressure (CoP), Forces (GRFs) and Moment (M) of forces as one moves across them, can be extracted.

The reliability of force-plate time series data with different test conditions has been proved and discussed by several researchers (Bauer et al., 2010; Golriz et al., 2012; Martina Mancini et al., 2012). The displacement of Center-of-Pressure (CoP), derived from the force-plate, is often used to characterize gait and balance (Hof et al., 2005; Ruhe et al., 2010). The CoP represents body motion in space as detected at the interface between the feet and the ground. The COP displacement time series represents the location of the (resultant) vertical Ground Reaction Force vector. This is used as a mean to quantify one’s balance: greater displacement of the CoP position could mean greater instability. Features such as Root-Mean-Square (RMS) of medial-lateral (M/L) and anterior-posterior (A/P) directions, Sway area, Axis length, etc. could be also extracted from time series.

Combining functional tests results with sensor-based measurement (e.g., force-plate) can provide valuable information for gait and balance analysis. Force-plate measurement provides more sensitive and precise measurement due to its application in the experimental environments and has decreased: test variability, subjectivity of the scoring system and sensitivity to small changes (M. Mancini & Horak, 2010a).

To draw a complete picture of the human’s gait and balance and investigate the motion’s kinetics and kinematics, dos Santos et al. used 42 markers, integrated with a 3-D motion capture system, on each subject’s full body. They recorded different parameters such as Ground Reaction Forces (GRF), Center of Pressure (CoP), Center of Gravity and participants’ joint angles through the combination of both motion capture and force-plate measurements. Using this approach, they managed to create a rich quantitative evaluation of human balance (dos Santos et al., 2017).

In Table 1, we summarize several studies that used combination of force-plate and other tools to record human’s biomechanical gait and balance features and investigated the effect of various factors on postural control. Each of the summarized studies have used various techniques to analyze human balance. Giovanini et al. classified balance characteristics of older adults as low or high risk of falling. They used the same dataset in this study (dos Santos & Duarte, 2016) which contains subjects’ CoP information in the form of time series signals. By applying six different Machine Learning (ML) techniques and extracting 34 temporal/spatial features from the subjects’ CoP, they showed high discriminative powers in the 60 seconds-time series. Among all ML methods in the study, the highest accuracy they achieved, was 64.9% by Random Forest (RF) classification method (Giovanini et al., 2018).

**Table 1.** Overview of studies on commonly used balance/gait metrics and different analyzing techniques

| Study | Research Objective | Used Features | Subjects | Measuring equipment | Analysis Method/tool | Study outcome | Max Acc |
| --- | --- | --- | --- | --- | --- | --- | --- |
| (Balestrucci et al., 2017) | To investigate the effect of dynamic visual cues on postural control | CoP sway parameters (e.g., M/L and A/P Standard deviation, sway area, etc.) | 44 healthy individuals | Force-plate | Statistical Analysis (e.g., t-test, ANOVA) | Gravity-congruent visual motion makes significantly reduced postural sway compared to gravity-incongruent one | N/A |
| (Fukuchi et al., 2017) | To examine the effect of running speed on gait-biomechanics variables | Kinetics and kinematics (e.g., cadence, stride length, joints angles, joints torque etc.) | 28 regular runners | 3D motion-capture system & an instrumented treadmill | Statistical Analysis (e.g., one-way ANOVA, Kruskal-Wallis) | Most of gait-biomechanics variables (other than foot-strike) are affected by running speed | N/A |
| (dos Santos et al., 2017) | To provide the subjects' full-body 3D kinematics and the GRFs in static balance, while changing support surface and visual cues | Human body's GRFs, CoPs, and 3D Kinematics (e.g., joints angles) | 27 young and 22 older individuals | 3D motion-capture system & force platform | Visual3D software and Python programming | Biomechanical characteristics were visualized and modeled (e.g., CoP & COG displacement at the A/P & M/L directions versus time, etc.) | N/A |
| (Giovannini et al., 2018) | To distinguish between different age groups using force-plate signals with different time-series duration | CoP's temporal, spectral and spatial features (e.g., root mean square (RMS) distance, sway path, mean frequency, etc.) | older adults (dos Santos & Duarte, 2016) healthy and post-stroke adult (Giovannini et al., 2018) | Force-plate | Statistical Analysis (e.g., Wilcoxon test, Mann-Whitney U-Test, etc.), Machine Learning (ML) (e.g., K-NN, SVM, MLP, RF, etc.) | For statistical analysis: optimal CoP duration varies based on group under study,<br>For ML analysis: 60 s duration CoP signals is more discriminative | 64.9% (RF) |
| (Reilly, 2019) | To classify fall-risk in older adults using effective feature selection methods (e.g., ReliefF, SAFE, etc.) | Time-domain and Frequency domain features extracted from CoP and Force signals | 163 young and older individuals (dos Santos & Duarte, 2016) | Force-plate | Machine Learning (e.g., SVM, K-NN, MLP, NB) | Feature selection methods identified the relevant features successfully, but were incapable of improving classifiers reliability while using only static balance measures | 80% (MLP) |
| (Montesinos et al., 2018) | To differentiate between fallers from non-fallers | CoP's Approximate entropy (ApEn) and sample entropy (SampEn) with different input parameters | 163 young and older individuals (dos Santos & Duarte, 2016) | Force-plate | Statistical Analysis (e.g., three-way ANOVA) | SampEn represents a better choice for the analysis of CoP time-series and to distinguish between groups | N/A |
| (Cetin & Bilgin, 2019) | To discriminate between young and aged groups | CoP and Forces' Standard Deviation (STD) | 163 young and older individuals (dos Santos & Duarte, 2016) | Force-plate | Machine Learning (e.g., SVM, K-NN, DT, LDA, etc.) | Force signals are better predictors than CoPs for group differentiation while using force-plate measures | 81.67% (SVM) |
| (Ren et al., 2020) | Using AI to evaluate different types of balance control subsystems determined by Mini-BESTest | CoP's extracted Traditional features (e.g., Mean of CoP displacement, etc.) and pixel-based features (e.g., skewness of gray levels for all pixels) | 163 young and older individuals (dos Santos & Duarte, 2016) | Force-plate | Machine Learning Regressors (e.g., RF, MLP, LR, etc.) | Low mean absolute errors (MAE) showed reliability of AI techniques for assessing balance control subsystems | N/A |

Reilly used (Reilly, 2019), the same open-source dataset (dos Santos & Duarte, 2016) to study risk of fall. At the first stage, they extracted 528 features in the time and frequency domain. Feature selection methods such as a filter-based feature selection algorithm called ReliefF, Self-adapting feature evaluation (SAFE), etc. were employed to find the optimal number of features for classification. With 18 features through the SAFE method, the experiment result showed the highest classification of accuracy of 80% using Multiple Layer Perceptron (MLP). The experiment was based on 73 older subjects (out of 163 total participants) of the dataset, who were either at the low or high risk of fall. Other classifiers used were Support Vector Machine (SVM), K-Nearest Neighbors (K-NN) and Naïve Bayes (NB) with performance range of 75 to77 % accuracy (Reilly, 2019).

Another work on the same data (dos Santos & Duarte, 2016), was done by Cetin and his colleague. In their work, they discriminated between young-aged groups using different machine learning techniques such as SVM, K-NN, Decision Tree (DT), Linear Discriminant Analysis (LDA), etc. Employing force signals of the dataset, the experimental result showed the highest classification accuracy of 81.67% by SVMs (Cetin & Bilgin, 2019).

In the above studies, statistical analysis and/or machine learning techniques, were employed by a domain expert who had related research experiences and extracted relevant features for fall risk detection or group discrimination. Santos and Duarte employed the ‘shallow’ Neural Networks approach to analyze the same data used in this study (dos Santos & Duarte, 2016). The results of their experiment showed the highest accuracy of up to 80%; suggesting that extracted features were not sufficient in improving classification results (Giovanini et al., 2018; Reilly, 2019).

Cetin and Bilgin, (Cetin & Bilgin, 2019) argued that achieving a relatively high classification performance is a function of only force signals and not the whole force-plate measures. However, the maximum accuracy of their study was still limited to 81.67%. These results agree with (Safuan et al., 2017) conclusion that extracting suitable features from gait/balance data and choosing the optimal feature extraction/selection methods are complicated and time-consuming processes. Their work showed that nonlinearities, high dimensionality, and high variability of time-series signals are major constrains.

As shown in Table 1, a Machine Learning (ML) algorithm or Statistical Method has the capability of interpreting the gait and balance biomechanical features and finding the relationship between variables. However as mentioned above, these methodologies typically require domains expertise and advanced experience levels which could be highly time and energy consuming and prone to error. Wang et al. used simple Neural Network (NN) along with Bayesian optimization algorithm to analyze human walking speed. The study involved feature extraction and feature selection stages for generating input features for a classifier (Wang et al., 2021).

Table 1 suggests that methodologies used by other scientists in research studies on falls are not adequate. It also shows that fall detection and human balance characterization can be a challenging task. Deep Learning (DL) models have been shown to have the capability of scanning a large dataset for relevant informative features without passing through the manual feature extraction stage of the traditional ML or statistical models (Horsak et al., 2020). Hoffman et al. took advantage of recurrent neural network (RNN) based on Long Short-Term Memory (LSTM) to investigate gait patterns of 42 participants. They experimented with a capacitive sensor floor to record walking kinematics of subjects. According to their result, using combination of sensor-based data and NN is a promising approach to be applied in in health and care (Hoffmann et al., 2021).

If adequately utilized, DL algorithms have the capability of providing automatic feature extraction, flexibility, and higher accuracy on huge amount of data. This is because unlike traditional ML algorithms, deep learnings can learn high-level features from high-dimensional data in a hierarchical manner which eliminates the need of domain expertise and the complex process of feature extractions (Goodfellow et al., 2016; Najafabadi et al., 2015). The supremacy of deep learning algorithms to reveal underlying patterns of big data without the need of performing prior feature extraction and feature selection processes, can result in considerably higher predictive powers, and more accurate results. Therefore, for a better result and balanced study, our experiment employed deep neural network algorithms to classify force-plate balance time-series signal to predict human’s balance impairment. We argue that this is a better approach.

Exploring this capability and superiority of the DL, we investigate the balance patterns in different people for an effective characterization, using the same open-source dataset used by researchers in table 1. Dataset was produced by Santos et al. (Santos & Duarte, 2016). The dataset contains the force-plate time-series signals recorded from 163 young and older individuals. Alongside the force-plate dataset there is another large meta data file composed of personal and other health characteristics of the subject such as age, gender, history of fall, background disease, functional tests’ results etc.

Although the available meta data is composed of different types of information about participants which could be used as binary classes, e.g. gender (male vs. female) or age group (young vs. old), vision (open vs. closed), surface (firm vs. foam) etc. we decided to discriminate the study subjects based on their Falls Efficacy Scale (FES) test score. FES is an international known test which measures fear of falling. It implies a person’s level of confidence for carrying out everyday activities (Kempen et al., 2008; Morgan et al., 2013).

We hypothesized that low, moderate, and high risk of falling can be characterized based on commonly-used balance metrics, such as those derived from the force-plate time series, in conjunction with deep learning algorithms: Recurrent Neural Network (RNN), Long-Short Time Memory (LSTM), One Dimensional Convolutional Neural Network (1D-CNN), and our developed One-One-One Deep Neural Network. Further, we hypothesized that our proposed One-One-One neural network is the most efficient neural network model in predicting human’s balance impairment using the force-plate time series signal.

## III. Materials and Methods

### A. Materials: Force-Plate and Meta Data

As explained above, the data used in this study is a publicly available dataset. It is accessible through both PhysioNet (DOI: 10.13026/ C2WW2W) and Figshare (DOI: 10.6084/ m9. figshare. 3394432) websites. The study in which the dataset was created, and its detailed description, can be found here (Santos & Duarte, 2016). The data was collected from 163 male and female participants of different ages (18-86 years old) and varied health conditions. Each participant completed a 1-2-hour study session, and all data were collected during this single session. To evaluate a person’s balance, he/she was asked to stand still on the force-plate for 60 seconds with arms at his/her sides and repeated it three times for four different test conditions.

Test conditions were defined as standing eyes-open on a firm surface, eyes-closed on a firm surface, eyes-open on a foam and finally eyes-closed on a foam. Both rigid surface and foam tests were done on a force-platform and the participants’ balance characteristics of Force, Moments of forces, and Centers of Pressure were measured. Participants were barefooted and looked at a 3 centimeters round target in front of them, located at a wall 3 meters away (dos Santos & Duarte, 2016; Santos & Duarte, 2016).

The force-plate data was acquired at 100Hz frequency. Since each person did twelve 60 seconds trials in this study, twelve .txt files, each composing of 6000 rows and 9 columns exist for each subject. Therefore, after merging data files, each person has 72000 row time-series signal and 9 feature columns specified as: Time, as well as tri-axial Force (Fx, Fy, Fz), and Moment (Mx, My, Mz), along the x, y, and z axes respectively (as shown in Figure 1), and Center of Pressure in ML and AP (i.e., CoPx, CoPy) recorded by the force-plate. Moment of force (Latash, 2012a), and can be determined by the following formulas:

**Figure. 1.**
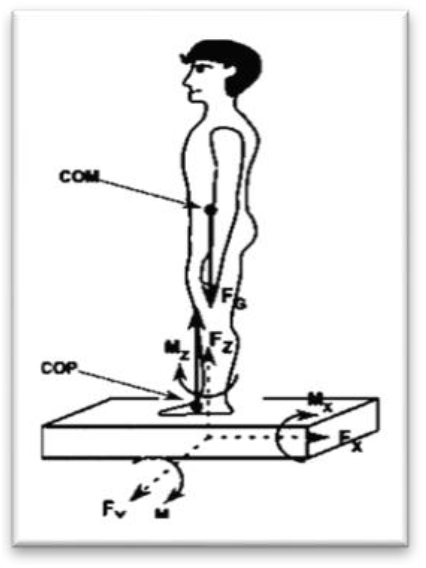
Measuring three components of the force vector (FX, FY, FZ) and three components of the moment-of-force vector (MX, MY, MZ) by force-plate; Retrieved from (Latash, 2012b)

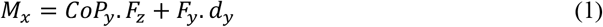

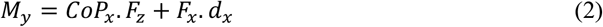

where M stands for Moment of force, F stands for force, dx and dy stand for the lever arm of the shear forces of F_x_ and F_y_ along the Z axis, respectively. Through those metrics, we can also determine Center of Pressure (CoP) with the help of the following simplified equations; (Latash, 2012b)

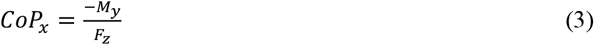

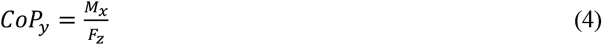

where CoPx and CoPy are the CoP coordinates along the X and Y axis, respectively.

Along with the main dataset which contains the subjects’ balance metrics, there is another information file, called BDSinfo, provided by the researchers. This supplementary file contains meta data about the subjects (e.g., vision, gender, height, BMI, illness, use of medication, number of falls, and disability). It also includes the results of some well-known conventional clinical evaluations such as Falls Efficacy Scale (FES), International Physical Activity Questionnaire (IPAQ), and Trail Making Test (TMT); these tests focus on assessing a specific parameter such as cognition level, concern of falling during different tasks and likelihood of fall and evaluate postural control of people and their stability (balance) while doing several static and/or dynamic motor tasks. (M. Mancini & Horak, 2010b). To understand the concept of each evaluation method and the whole meta data, reading the main source (Santos & Duarte, 2016) is highly recommended.

Due to its comprehensive information, the meta data is very resourceful for categorizing human subjects. In other words, each of the columns of this dataset can be used as a proper label for classification purposes. The available meta data is composed of different types of information about participants which could be used as binary classes, e.g., gender (male vs. female) or age group (young vs. old), vision (open vs. closed), surface (firm vs. foam), etc. However, we decided to discriminate the study subjects based on their Falls Efficacy Scale (FES) test score. The Falls Efficacy Scale International test (FES) is a well-known evaluation methodology used for individuals with vestibular or balance dysfunction to quantify his/her fear of falling. Participants answer to the questions about some of their regular daily activities and specify how concerned they are about the possibility of falling. Reliability and validity of FES to measure concerns of falling in people with imbalance and postural control deficits have been explained by several researchers (Kempen et al., 2008; Morgan et al., 2013; Yardley et al., 2005).

### B. Data Preparation

The code was written in Python 3.7.3. As the first step, we merged all .txt files recorded with the force-plate for all subjects. As described earlier, time series were recorded by 100 Hz frequency which produced 6000 rows for each 60 second trial. Since each of the 163 subjects completed the 12 trials, therefore we had 1956 .txt files. However, 26 of these .txt files are missing for five subjects who were not able to complete the most challenging exercises. Consequently, there are 11 580 000 rows of data recorded for all subjects.

### C. Label Selection

In this study, a supervised learning approach was used for the design, development, and evaluation of our learning algorithm. Using this approach, suppose we have a set of N training examples {(*x*_1_, *y*_1_), … (*x*_*N*_ *y*_*N*_)}, where *x*_*i*_ and *y*_*i*_ are the *i*^*th*^ feature vector and label respectively. The goal of the learning algorithm is to choose the optimal function *g: X → Y*, where *X* and *Y* are the input and output states respectively, function g belonging to a hypothesis space G. This hypothesis space represents all possible functions of G.

We chose 14 columns of meta data available at (dos Santos & Duarte, 2016) containing the most important information about participants and added those features to our main force-plate dataset. As discussed in section III, the main balance metrics recorded by force-plate were Time, Force in x, y and z directions (Fx, Fy, Fz), Moment of force in x, y and z directions (Mx, My, Mz), Center of Pressure in x and y directions (CoPx and CoPy). Thus, our feature size increased to 23 after adding the 14 columns of vision, surface, age group, gender, Bone Mineral Density (BMI), illness, Number of medications, disability, falls12month, IPAQ_S, TMT_timeA, TMT_timeB, Best_T, FES_S of meta data. IPAQ is an International Physical Activity Questionnaire Short Version test (IPAQ) which measures health-related physical activity (PA) in populations (Hagströmer et al., 2006). TMT is abbreviation of Trail Making Test which screens dementia by assessing cognition level in two different parts A and B, and the goal is to complete the tests accurately and as quickly as possible (Salthouse, 2011). Mini Balance Evaluation Systems Test (Mini-BESTest) predicts the likelihood of fall (Yingyongyudha et al., 2016).

In this research, we merged the force plate and meta data file to create our dataset and used the Falls Efficacy Scale (FES) as the label. The proposed classifier in this study is based on the FES test result which provides three different classes: low, moderate and high fear of falling. The reliability and convergent validity of this test has been proven by different researchers (Kempen et al., 2008; Morgan et al., 2013). Using FES test results, subjects in this study are categorized into low fear (scores 2-9), moderate fear (9-13), and high fear (>14) of falling. We aim to distinguish between them using deep neural networks.

### D. Neural Network

Inspired by the human’s brain, Neural Network (NN) is a set of interconnected artificial neurons which recognize underlying relationships between datapoints. Each neuron or perceptron is a mathematical function that takes the input data from the input layer (x_1_, x_2_…), multiply them by a weight (w_1_, w_2_…), and add a bias (b) to the weighted inputs (hidden layer). To introduce nonlinearity to the network, the result is then passed through an activation function (f). Through feeding neurons in a forward way and consecutive computational processes, patterns are recognized by the network and the output is predicted (output layer). To achieve the best estimation of desired outputs, the system adjusts the weights and bias continuously using backpropagation of error (B. Wankhede, 2014; Hecht-Nielsen, 1989). Figure 2 demonstrates a simple architecture of neural network composed of three layers. For a more complex decisions, networks consist of several hidden layers and neurons are recommended.

**Figure. 2.**
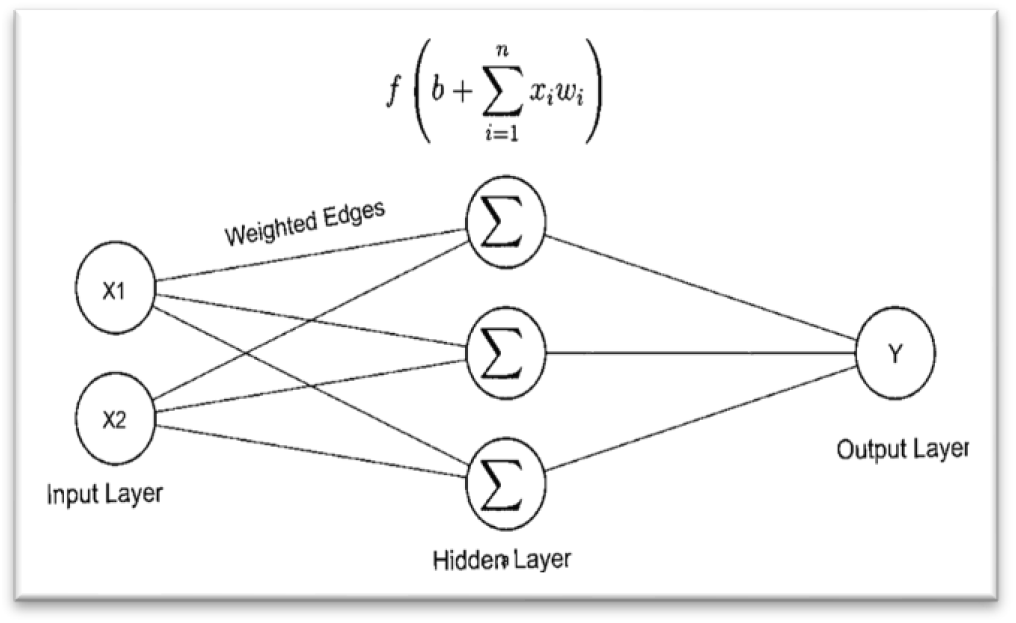
A 3-layer neural network architecture

### E. Experimental design

Figure 3 summarizes the flowchart of our experimental design. As shown in the diagram, our datasets comprise of two separate files: force-plate time series and meta data files. Thus, the first step was merging the two files. The time vector was removed from the input data since it just represented the elapsed time and did not contain any other information about subjects’ balance characteristics. Samples were randomly selected. Random sample selection reduces skewness, class imbalance and overfitting. It is also possible to generate several samples. This strategy has the advantage of repeatability of experiment. Besides being time and space efficient, random sampling provides an equal chance for each datapoint to be selected. Thus, it improves the dependability, reliability, and efficiency of the deep learning models (Gonçalves et al., 2012).

**Figure. 3.**
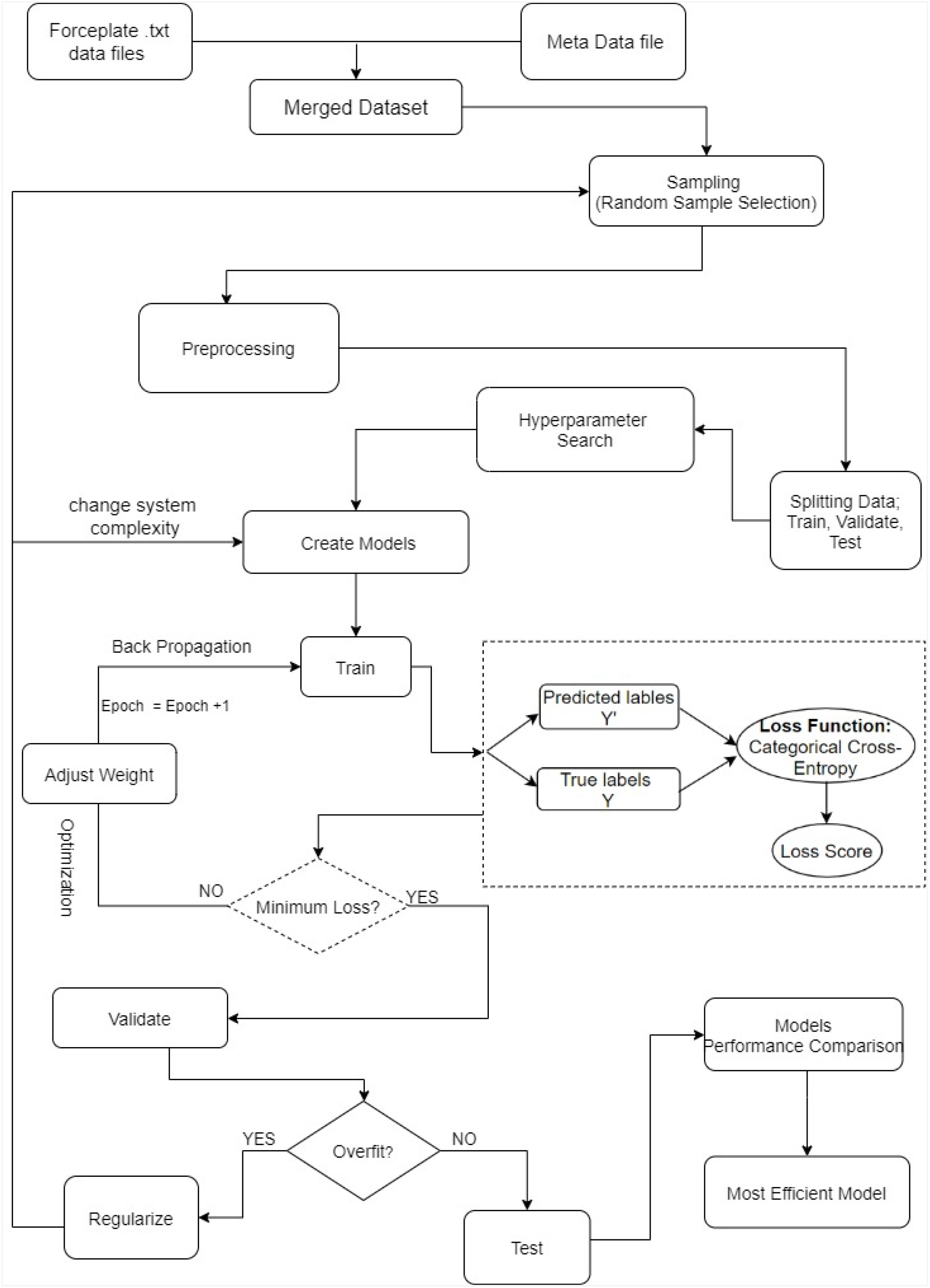
System Architectural flowchart

Features were scaled, and labels were encoded in the preprocessing stage. Data was split into 70% train and 30% test data. Different deep neural network models were selected for the experiment. Optimal hyperparameters were calculated for each model and data was fed into the learning algorithms. Getting the exact values of the hyperparameters are critical because these parameters guide the training process; hyperparameters have a strong influence on the performance of a deep learning model. In other words, “wrong” choices of hyperparameter values are most likely to produce poor performance of any learning algorithm (Liu et al., 2006).

Unlike other parameters which are learned in the training process, values of hyperparameters are set prior to the training. In most deep learning applications, some important hyperparameters include number of epochs, batch size, number of hidden layers, optimizers, activation function, regularizations etc. In this study, we used the Grid Search approach of hyperparameter search for the optimal values of required hyperparameters. Therefore, our systems’ structure was based on the values obtained from this tuning process. Building the models’ architecture using the optimal hyperparameters values reduce the risk of overfitting thereby improving generalizability.

Deep learning algorithms categorize data through layers of computational neurons. Each neuron has a certain weight and works with an activation function, such as the “sigmoid”. In each iteration, the output is determined by the system based on the learnt information, error is calculated and fed back to the system using the back-propagation techniques. For an optimal performance, difference between the expected label and the predicted outcome should be minimal. But in most experiments, it takes more than one iteration to achieve the minimum error. Therefore, weights are adjusted at each iteration. This process is called optimization and continues until the maximum accuracy is achieved with the lowest error. Each neural network model has a capacity or complexity which is defined by its structure (number of weights) and parameters (values of weights). Therefore, to avoid/fix overfitting, the system’s complexity can be modified by various regularization techniques such as: Weight Regularization, Activity Regularization, Weight constraints, Dropout, Noise and Early Stopping (Brownlee, n.d.; Ying, 2019).

For validation purposes, 20% of the training data was assigned for validation. As an evidence of generalizability, we expect a consistent overlap behavior between the training and validation learning curves. However, optimal consistency is not guaranteed. This can be due to the problem of overfitting. Overfitting occurs when a model learns the training dataset too well but does not perform well on the validation data.

The loss function computation for the proposed neural network models is “Cross-entropy”. Loss functions measures the difference between actual value of target variables and how robust the system works. To determine the loss score through “Cross-entropy” the following equation is used:

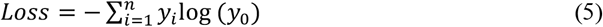

where *n* is the number of training samples, *y*_*i*_ is the predicted value and *y*_*0*_ is the actual value of label. A small loss value represents good performance of our used classifier (Ho & Wookey, 2020). After each iteration, the loss score is calculated. Then a selected optimizer, was used to update the weights in a back-propagation manner to minimize the loss and increase accuracy. The model was trained, optimized, and validated continuously. The model training stops when the loss of validation set stops decreasing.

The reliability, effectiveness and repeatability of our experiment was demonstrated with multiple neural network models. We compared the performance of the models to determine the most efficient one for predicting human’s fall-risk using the force-plate signal dataset. Performance comparison of the models were based on accuracy, losses, and number of iterations to attain optimization. We also compared precision and specificity. For easy comparison, experimental results are shown in graphical and tabular formats.

#### E-II. Multiple Layer Perceptron (MLP)

The Multiple Layer Perceptron (or MLP) is a variant of the deep learning neural network family. It consists of an input layer, two or more hidden layers and an output layer.

For D input features x_1_, …., x_D_ and M numbers of nodes j, *j =* 1,…, *M*. The M linear combinations of the input features at the Hidden Layer L1 (Hidden Layer L1 is the second layer of the network) is given as;

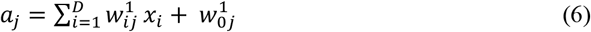

Where 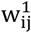 is the weight parameters connecting the *i*^*th*^ node at the first layer to the *j*^*th*^ node at the second layer.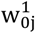 is the bias parameter a_j_ is then transformed using a differentiable non-linear activation function h such as the “sigmoid”;

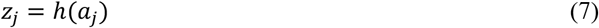

Z_j_ is passed to Hidden Layer L2 (Hidden Layer L2 is the third layer of the network)

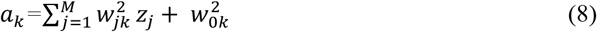

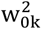 is the bias parameter for L2.

a_k_, is transformed using an activation function.

The process continues.

Combining equations 6 and 8, the sets of weight and bias parameters can be represented as a vector W, and the input features as a vector X.

Mathematically,

The output *y*_*k*_ (*X,W*) =

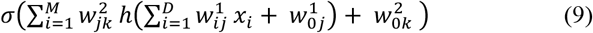

Equation 9 suggests that the output y_k_ is a nonlinear function of sets of input features {x_i_} and adjustable parameters vector *W*. We can reduce the size of equation 9 by defining a variable *x*_*0*_ such that its value equals 1, thereby the bias parameter is represented in the weight parameters vector W. Then, equation 9 becomes;

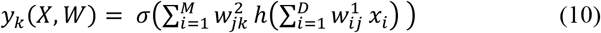

In general, given an output label y with input vector X, an MLP finds the best function *f* mapping *y= f(X, W)* while learning the optimal value of the parameter W.

MLPs are also called feedforward neural networks because of the flow of information from the input to the output via the intermediate layers. It exhibits a directed acyclic architectural graph represented as a chain structural function of the form;

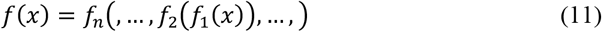

Where *f*_*n*_, *f*_2_ and *f*_1_ are the *n*^*th*^, second- and first-layers non-linear functions, respectively.

An example of MLP comprising of an input layer, two hidden layers and an output layer is shown in figure 4. As shown, the architectural layout of an MLP depicts that all layers in the network are fully connected. This implies that all nodes in the middle layer are connected to all nodes in the next and previous layers. All nodes in the input and output layers are connected to all nodes in the next and previous layers respectively. For example, as shown in the diagram, each node in layer 1 is connected to all the nodes in layer 2, all nodes in layer 2 are also connected to all nodes in layer 3, the process continues till the output layer.

**Figure. 4.**
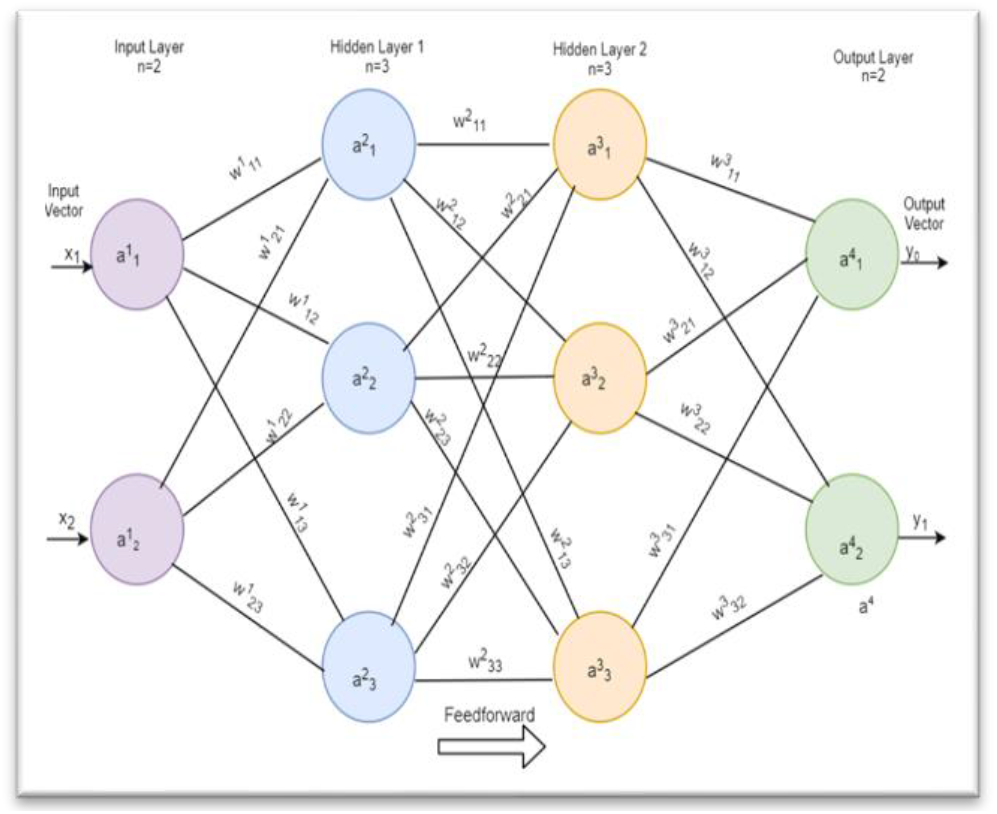
A Multiple Layer Perceptron consisting of an input layer with 2 input features x1 and x2, and 2 hidden layers each with 3 neurons and an output layer with 2 neurons

For a complex MLP network, layers can be in hundreds or thousands. The input layer is considered as the first layer, hidden layer 1 as the second layer, hidden layer 2 as the third layer, etc. Edges between nodes from one layer to the next are denoted as w_ij_^n^, where *n* is the layer number, *ij* is the weighted edges connecting the *j*^*th*^ node in the *n*^*th*^ layer to the *i*^*th*^ node in the *(n + 1)* ^*th*^ layer. For example, the weight connecting the second node in the first layer (input layer) to the third node in the second layer (hidden layer 1) is denoted as w^1^23. Using the annotated diagram in figure 4, the above relationship can be represented in matrix form for easy computation. The input features are vectorized:

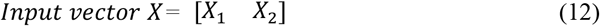

The weight matrix at Hidden Layer L1 is computed and multiplied by the input vector. Result sent through an activation function σ (e.g sigmoid) for non-linearization.

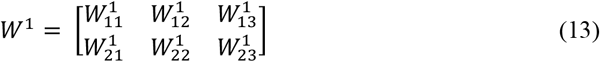

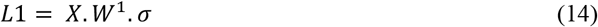

The weight matrix at Hidden Layer L2 is computed

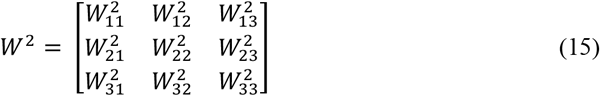

The output of Hidden Layer L1 is passed to the Hidden Layer L2 and multiplied by the activation function

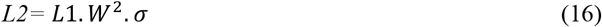

Depending on the number of hidden layers, the chain continues, and the final layer is passed to the output layer

The architectural arrangement of an MLP shows that it has the mechanism of performing complex computation, however, at a high cost. Furthermore, it suggests that an MLP is a global connected neural network, lacking the capability of exploiting a spatial or temporal representation of a dataset. This high space and time complexity of an MLP is its major drawback.

Since our dataset is spatial as well as temporal, our models are built on the flavors of convolutional and recurrent neural networks. Therefore, the experimental design comprises of the following deep learning models: 1D-Convolutional Neural Networks (CNN), Vanilla Recurrent Neural Network (VRNN), Long-Short-term Memory (LSTM) and a proposed One-One-One Neural Networks. The next section discusses the architectures of each of the four models.

#### E-III. 1D Convolutional Neural Network (CNN) Model

Convolutional Neural Network (CNN) is a popular method in deep learning. Convolutional layers are made of basic structures known as feature detectors or kernels. Unlike the MPL where weights are assigned directly to each feature, CNN is based on kernel feature engineering. Each kernel is a matrix of integers sliding on the input data as a filter to detect necessary informative features for an efficient representation and characterization of the dataset. There is an abundant application of CNN to images and videos. Poma et al. investigated optimization of CNNs using Fuzzy Gravitational Search Algorithm method (FGSA) for pattern recognition and image classification (Poma, Melin, González, & Martinez, 2020; Poma, Melin, González, & Martínez, 2020). However, it has been proven that 1D-CNN can be applied efficiently for time series signals analysis (Jiang et al., 2019).

As shown in figure 5, the architecture of our 1D CNN model comprises of first two consecutive convolutional layers, follows by a max pooling layer. For this experiment, using two consecutive convolutional layers at the initial stage of learning before max pooling has the advantage of preserving the true spatial representation of the dataset. Unlike images and videos that may need more convolutional layers, we limited the consecutive convolutional layers to only 2. Intuitively, information lost is minimized. This is logical because we are experimenting with force-plate time series signal. There are 32 filters in each of the convolutional layers.

**Figure. 5.**
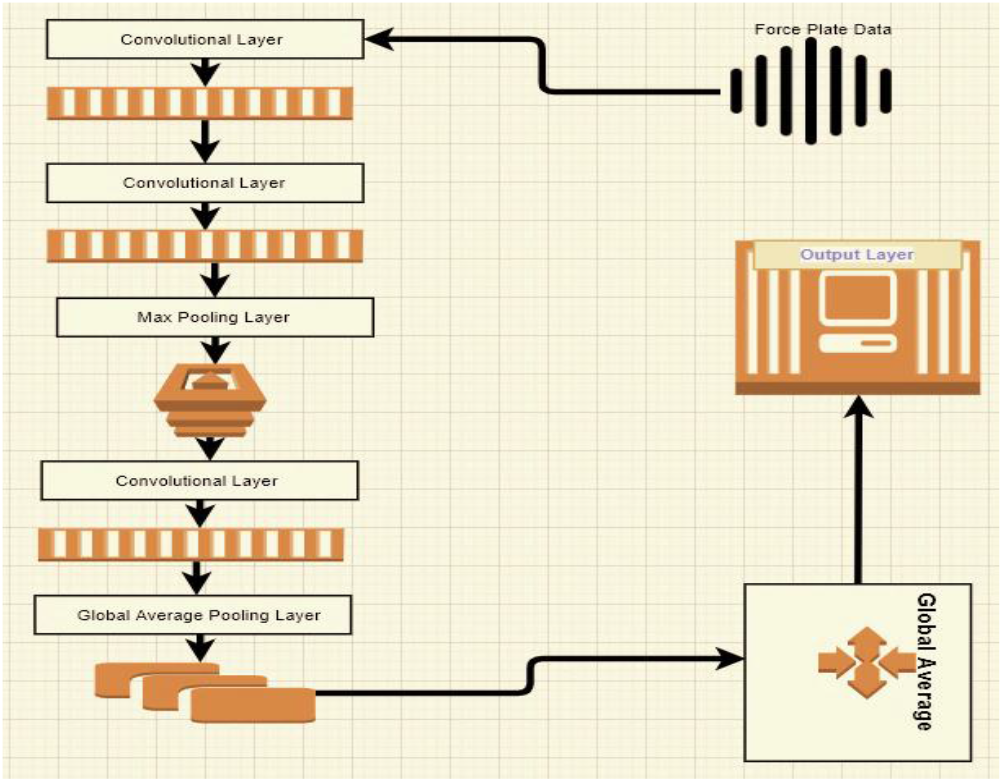
Schematic architectural diagram of the 1D-CNN used in this study, composed of three convolutional layers, Max Pooling and Global Average Pooling

From a mathematical viewpoint, convolution is a dot product of input and kernel functions which ultimately results in convolved features, also known as feature or activation maps (Wu, 2017). There is an activation function called Rectified Linear Units (RELU) on top of each convolutional step. This adds non-linearity to the extracted features and improves the discriminatory capability of the system (Kuo, 2016).

Features were extracted by the 1D-Convolution Network from each segment of force-plate data using the following equation:

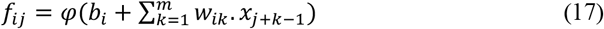

In equation (17), *f_ij_* is the extracted features vector from the jth neuron of the ith filter in the hidden layer, φ is the activation function which was assigned as “RELU” in this work, *b_i_* is the i-th filter corresponding overall bias, *w*_*ik*_ the featuring weight matrix, and *x*_*j+k-1*_ represents the input signals vector (Jiang et al., 2019). “RELU” provides the non-linear transformation of the input data for a better hypothesis space generated from its deeper representation. Without the non-linear activation function, the model will only be limited to the dot product and addition linear operations. “RELU” outputs the input number if it is greater than zero otherwise it outputs zero. Mathematically, “RELU” is represented as; g*(z)=max (0, z)*, where z is the input number.

At the pooling layer, we reduced the variance and computational complexity of the dataset. Pooling is the dimensionality reduction path of the 1D CNN model. Pooling operation can be minimum, average, or maximum, summarizing the least, average, and most activated features in each patch of the feature map respectively (Yamashita et al., 2018). The most popular approaches are the maximum and average pooling, minimum pooling is rarely used. In this study, we used max pooling because unlike the average pooling, max pooling provides the maximum presence of a feature. Therefore, max pooling tends to preserve the most valuable information. We used a 2 by 2 window with the stride of 2 for the max pooling layer. After the pooling layer we have another convolutional layer and finally a global average layer. The third convolutional layer has 64 filters. Each of the filters has a kernels size of 1*3.

The conventional approach is feeding information extracted through the convolution and pooling layers into a fully connected dense layer. This completes the process of data characterization. However, for this experiment, we did not use a fully connected layer. Instead, the output of the last convolution layer is fed into a global average pooling layer. The global average pooling layer computes the average value of each feature map. Computed averages are sent directly into the “softmax” layer for classification. It has been shown that global average pooling is less prone to overfitting and more robust to spatial data translations when compared with the fully connected layer (Lin et al., 2013). The output layer is made of 3 neurons and “softmax” activation function. To get the best out of our network, we also performed Grid Search and determined the optimal value for the hyperparameters.

#### E-IV. Recurrent Neural Network (RNN) and Long Short-Term Memory Models

We continued our investigation by training a Vanilla Recurrent neural networks (RNN) deep learning model. RNNs are variants of Neural Networks that process input data through number of layers in which the output of each step is dependent on previous computations. In other words, RNNs have a short-term memory which saves the calculated information and uses it for further analysis in the next layer. In fact, RNNs are several copies of the same structure consist of loops which allow the information to persist. This chain-like nature makes RNNs as powerful tools for different applications such as speech recognition, language processing, translation, image captioning, etc. The problem with RNNs is their short memory and looking at just the recent information disables the model to look back longer. A proposed strategy for solving this drawback is using a long short-term memory network (LSTM).

Figure 6 shows the architecture of an LSTM. As shown in figure 6, each memory cell is composed of input, forget and output gates *f*_*t*_, *i*_*t*_ *and o*_*t*_ respectively. The “sigmoid” function *σ* in the forget gate ‘looks’ at the previous state *h*_*t*-1_ and current input *x(t)* and decides what information should be discarded at the forget gate. Using “sigmoid” and “tanh” functions, the input gate decides about which input values should be. Finally, the output gate uses a “sigmoid” function to decide what parts of the cell we are going to output (*O*_*t*_) and then employs a “tanh” function which is multiplied by the output of “sigmoid” and gives weights to the values based on their level of importance *(h*_*t*_).

**Figure. 6.**
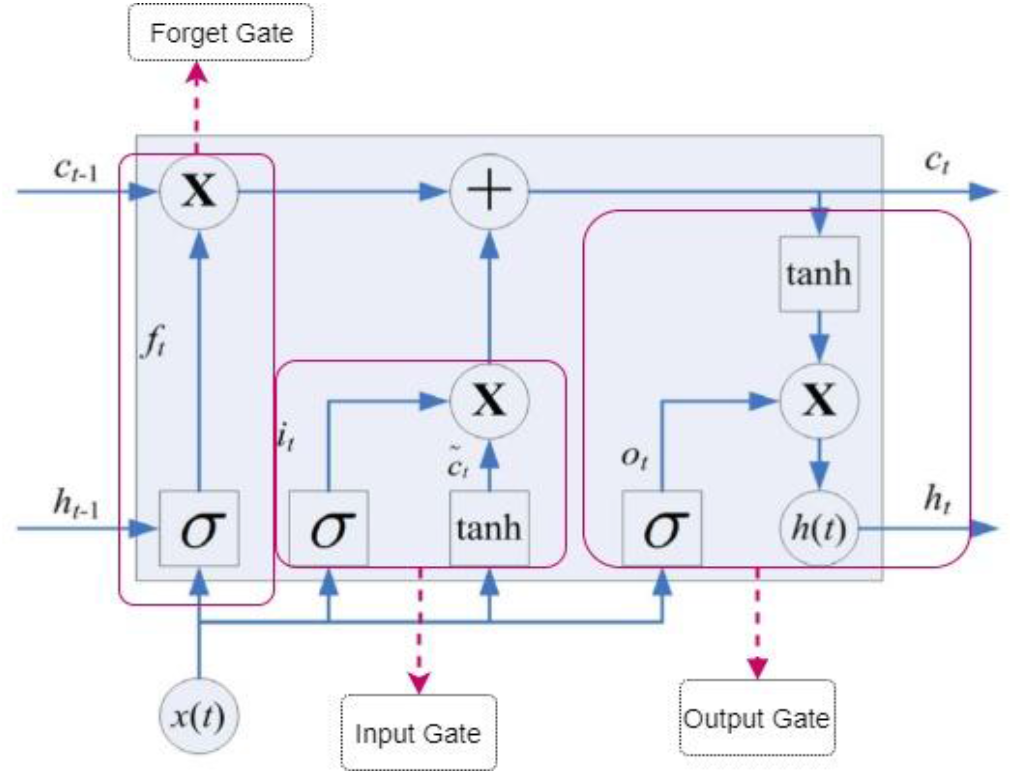
Memory cell of a long short-term memory network (LSTM); (Olah, n.d.; Staudemeyer & Morris, 2019)

The mathematical formulation of an LSTM is shown below;

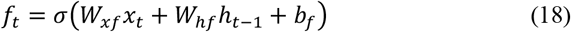

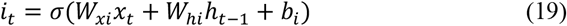

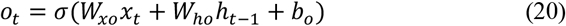

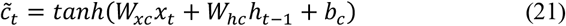

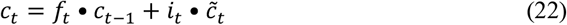

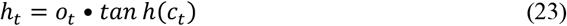

As shown in equations (18 to 23) *W_f_ W*_*i*_ *and W*_*o*_ are the weight matrices at the forget, input and output gates respectively, while *b*_*f*_, *b*_*i*_ and *b*_*c*_ are the biases in the same order. *f*_*t*_ and *i*_*t*_ are the activation vectors for the forget and input vectors. Detailed Mathematical description of LSTM and its architecture can be found at (Olah, n.d.; Staudemeyer & Morris, 2019; Zhou et al., 2015).

Due to this advanced hierarchical manner, it was hypothesized that better result is achievable using LSTM system rather than RNN. In fact, it is assumed that by using LSTMs we can overcome the Vanishing Gradient problem of RNNs and get a more accurate classification at lower computational cost (Tiwang, Oladunni & Xu. 2019). To test this hypothesis, we employed both the simple RNN and LSTM models. RNN was designed with 64 units along with a dense layer of 3 neurons and “softmax” activation. The LSTM architecture was designed with one LSTM layer of 256 units along with two dense layers each made of 128 neurons and an output layer of 3 neurons with “softmax” activation.

#### E-V. Proposed Model – The One-One-One Neural Network for Human’s Balance Impairment Prediction

The performance of models built with CNN, RNN and LSTM architectures described in sections E-III and E-IV proved to be inadequate; i) 1D-CNN attained 99.3 % accuracy at the 50^th^ epoch, ii) RNN and LSTM learnt faster when compared with the 1D-CNN but achieved accuracies of 96.9% and 98.3% respectively. Therefore, we explored a combination of 1D-CNN, LSTM and the Dense.

*We hypothesized that the proposed one-one-one neural network is the most efficient neural network model in predicting human’s balance impairment using the force-plate time series signal*. The optimization of the proposed model is based on; i) random sampling for data selection, ii) architectural simplicity and minimum complexity approach, and iii) Exhaustive search technique of hyper parameters’ values using the grid searching methodology. The proposed optimization approach is in line with Occam razor principle of parsimony and plurality which have been shown to improve generalization (Clark, n.d.). It is also in line with the Isaac Newton rule 1 of scientific reasoning (Four Rules of Scientific Reasoning from Principia Mathematica, n.d.). Furthermore, it agrees with Minimum Description Length (MDL) principle. MDL is a trade-off between the complexity of the model and the goodness of fit. Overly complex modeling has been linked to overfitting (Grünwald, 2007).

##### i. Random Sampling

As described in section III, our dataset comprised of 11 580 000 rows recorded for all subjects. A Microsoft Surface Laptop 2, Core i7 processor, 8 GB RAM was available for the experiment. It took 24 hours just to merge the files. Since merging data files was highly time-expensive on the laptop, intuitively, using this device for running deep learning algorithms with large number of epochs could be dramatically more expensive, laborious and may lead to an unsuccessful experimental outcome. The question here is; *do we need 11M records of the dataset to build the proposed model?* If the answer to this question is no, then the next question is; *can we reduce the number of records and still maintain a balanced class for a reliable experiment?* To answer these questions, we used a random sampling approach. A code was written which randomly selected a subset of the dataset. The result of our experiment shows the effectiveness of the approach.

##### ii. Architectural Simplicity and Minimum Complexity of the Proposed Neural Network Architecture

To test our hypothesis and investigate the efficiency of architectural simplicity and minimum complexity approach, we designed, developed, and evaluated three models from high to low complexity level: 1) One-One-Three, 2) One-One-Two, and 3) One-One-One neural networks. The first and second layers of all the three models have one 1D-CNN and one LSTM comprising of 64 filters with a kernel size of 1*3 and 256 units, respectively. However, we designed different dense layers for each architecture. In One-One-Three, there are three dense layers of 128, 64 and 32 neurons. In One-One-Two, there are two dense layers of 128 and 64 neurons. The proposed One-One-One deep neural network comprises of only one dense layer of 128 neurons.

As shown in figure 7, feature extraction was done at the convolutional layer. The filters ‘looks’ through the window for a true characterization of the dataset. Unlike the CNN architecture at section E-III where we used two consecutive convolutional layers at the beginning, here we used only one layer. Using one layer reduces complexity.

**Figure. 7.**
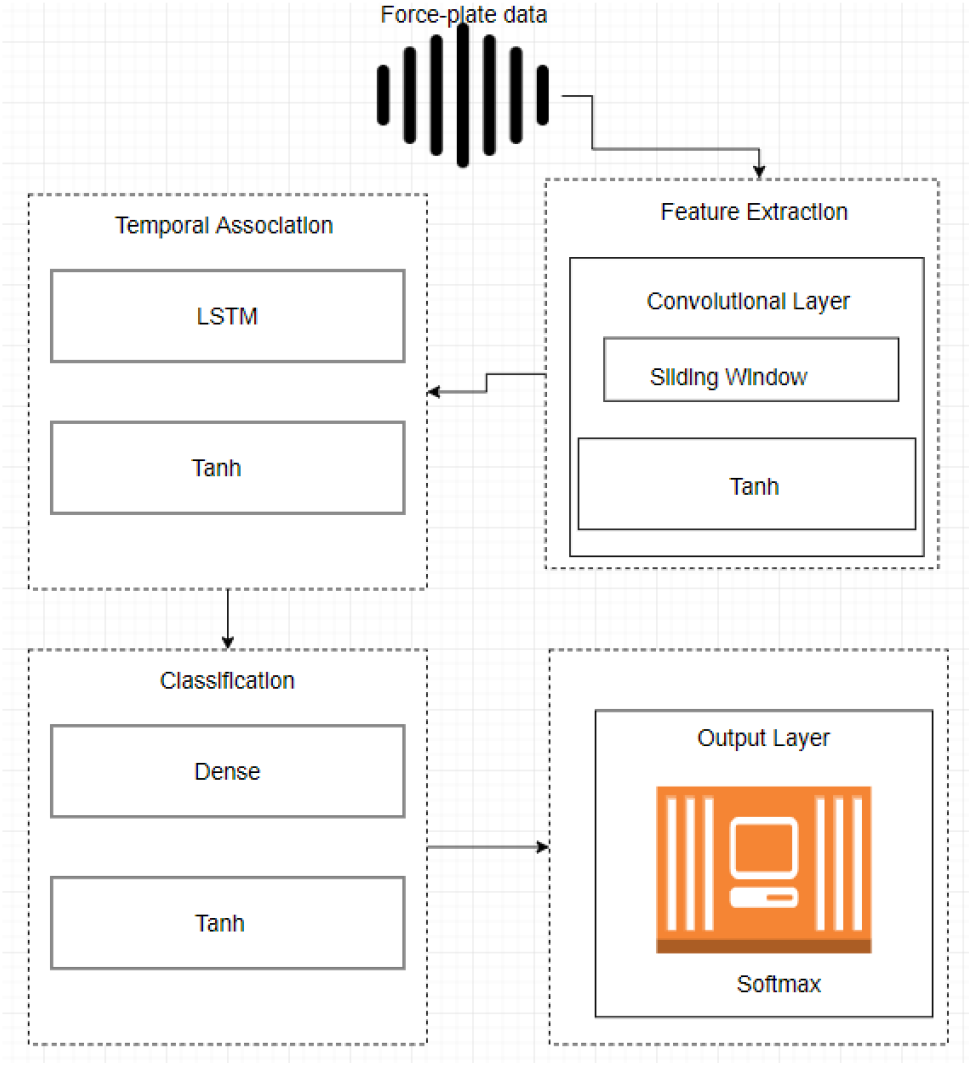
Proposed One-One-One Neural Networks Architecture; composed of one 1D-convolutional layer, one LSTM layer, and one dense layer

After the 1D-convolutional layer, we employed one LSTM layer composed of 256 units. LSTM provides information about the temporal associations of the features extracted at the Convolutional layer. “Tanh” activation function was used for non-linearity. Output of the LSTM was passed to a fully dense connected layer. The output layer comprises of 3 neurons with the “softmax” activation function.

##### iii. Hyperparameter Tuning

Building an efficient deep learning model is a very challenging task. One of these challenges is the selection of hyperparameters’ values. Choice of hyperparameters’ values affect the generalization capability and the overall classifier performance (Bergstra & Bengio, 2012; Liu et al., 2006). Figures 8 and 9 show the accuracy and loss graph of an un-tuned classifier using some randomly selected hyper-parameters’ values.

**Figure. 8.**
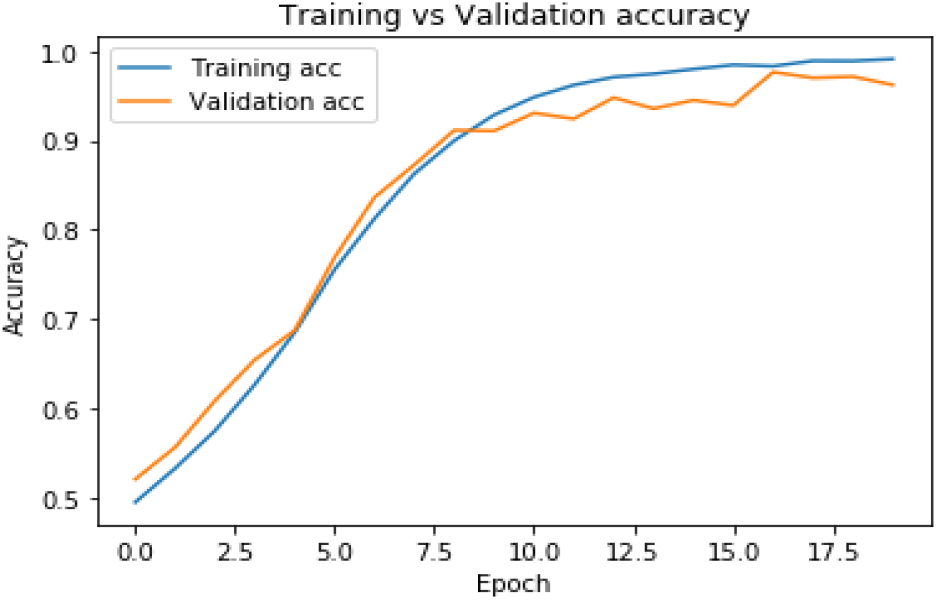
Training vs. Validation Accuracy of an un-tuned classifier

**Figure. 9.**
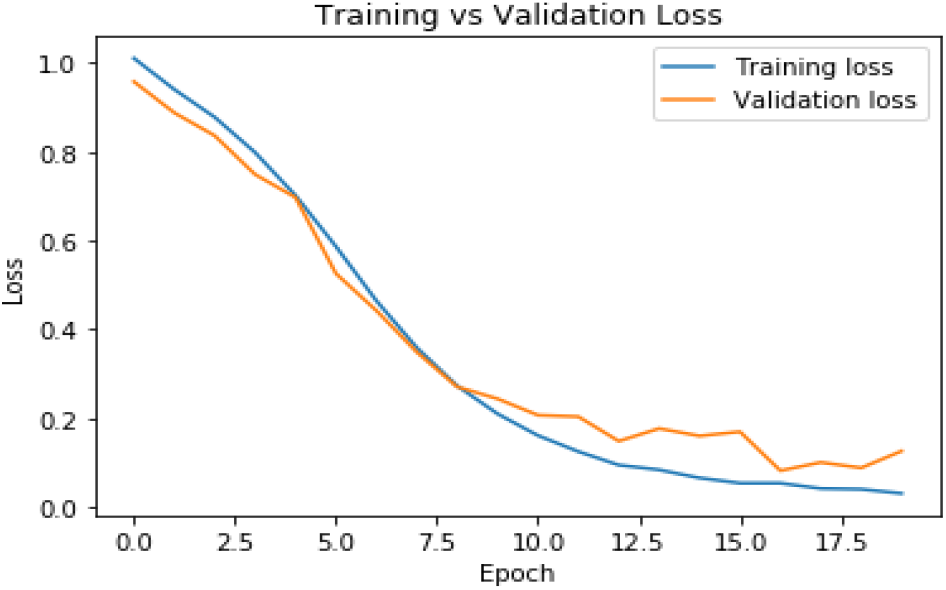
Training vs. Validation Loss of an un-tuned classifier

Looking at the figures, it is obvious that the training and validation graphs at some point show convergence, however, after a while they became unstable. Therefore, choosing the appropriate hyperparameters’ values is a crucial step in this experiment. The question here is, *how do we choose the best values of hyperparameters for a reliable and optimal performance of the proposed model?* We answered this question by computing a Grid Search of hyperparameters’ values for the experiment. Random search and manual selection are other popular approaches of choosing the values of hyperparameters. Grid Search is our preferred choice because it is the most exhaustive search approach when compared with the manual and random search. Successful implementation of the manual selection in a reasonable time depends to a large extent on the experience of the researcher. On the other hand, time and space constrain of the random search makes its iterations non-exhaustive. Grid Search produces better accuracy at a higher computational cost.

We imported GridSearchCV from the sklearn library in python programming environment and defined our desired model at the estimator component of the GridSearchCV module. We did this process for the 1D CNN, RNN, LSTM and the proposed One-One-One Neural Networks. The best values of each hyperparameter were determined. Through Grid Search we obtained the best values for the number of epochs, batch size, activation function, optimizer etc.

## IV. Results

We implemented our experiment based on the architectural designs of section E explained in methodology. 1D CNN, VANILLA RNN, LSTM and the proposed One-One-One Neural Networks models were trained, tested, and evaluated. Results of each model is discussed in this section.

### A. Performance Evaluation

To evaluate our models’ performance, we used confusion matrix and its four different outcomes, namely true positives (TP), true negatives (TN), false positives (FP), and false negatives (FN) (Table 2). These metrics are produced as a result of classification predictions and are employed to evaluate the model’s performance by calculating its sensitivity, precision and accuracy through the following formulas:

**Table 2.** Summary of experimental results

| Model | Confusion matrix |  |  | Precision (%) | Sensitivity (%) | Acc (%) |
| --- | --- | --- | --- | --- | --- | --- |
| CNN | 610 | 1 | 5 | 99.3 | 99 | 99.3 |
|  | 1 | 977 | 2 |  |  |  |
|  | 8 | 4 | 1392 |  |  |  |
| RNN | 595 | 9 | 23 | 96.6 | 96.6 | 96.9 |
|  | 4 | 935 | 17 |  |  |  |
|  | 18 | 20 | 1379 |  |  |  |
| LSTM | 662 | 7 | 6 | 98.3 | 98.6 | 98.3 |
|  | 0 | 939 | 19 |  |  |  |
|  | 1 | 17 | 1349 |  |  |  |
| One-One-Three NN | 585 | 2 | 1 | 99.3 | 99.6 | 99.5 |
|  | 2 | 999 | 8 |  |  |  |
|  | 0 | 2 | 1401 |  |  |  |
| One-One-Two NN | 604 | 0 | 1 | 100 | 99.6 | 99.7 |
|  | 0 | 960 | 1 |  |  |  |
|  | 0 | 5 | 1429 |  |  |  |
| Proposed One-One-One NN | 631 | 0 | 0 | 100 | 100 | 99.9 |
|  | 0 | 998 | 2 |  |  |  |
|  | 0 | 1 | 1368 |  |  |  |

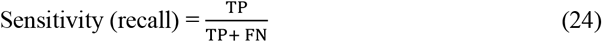

Sensitivity is the ratio of truly predicted labels belonging to a class to all samples that truly belong to that class. A higher value of sensitivity represents higher value of true positive and thus lower value of false negative predictions.

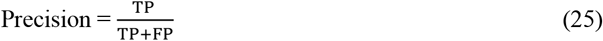

Precision is the ratio of truly predicted labels belonging to a class to all samples that were predicted to belong to that class by the classifier. Precision shows the relevance of positive detections.

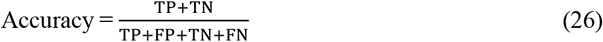

Accuracy is the ratio of correct samples predictions to total number of all predictions and shows the general model’s performance in terms of correct classification. Determined high value of all used metrics can be representative of the model’s high dependability and discriminative power.

### B. CNN Model

As discussed earlier, the model used two 1-dimensional convolutional layers, Max Pooling and Global Average Pooling. Since we are working with numerical force-plate time-series signal here, and not pictures, using 1-D Convolutional Neural Network turns out to be more appropriate. Figures 10 (a & b) show the accuracy and loss results of our designed model along with validation results. As it is seen in the graphs, there is a high consistency between accuracy and loss of both training and validation data.

**Figure. 10.**
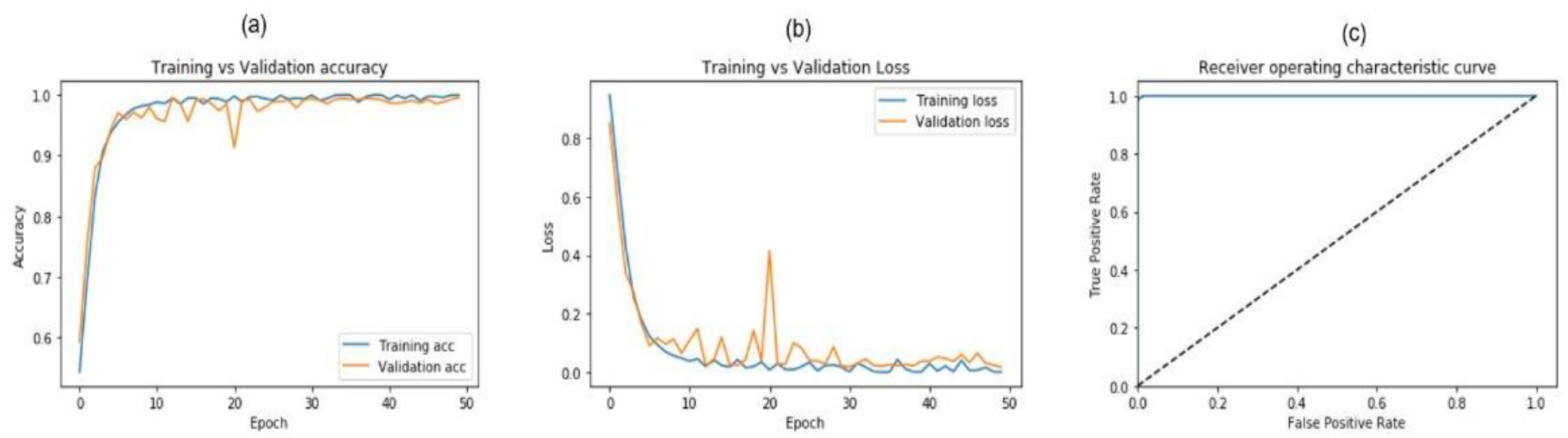
CNN classifier’s Training vs. Validation (a) Accuracy (b) loss (c) Receiver operating characteristics.

The highest classification accuracy of 0.993 was achieved at the 50th epoch. The reliable performance of the model in predicting true labels can be also observed through the receiver operating characteristics graph shown in figure 10 (c).

### C. RNN and LSTM Models

We also trained and evaluated the RNN and LSTM models. As discussed earlier, for the RNN model, we used a simple RNN. The number of units and neurons were defined as 64 and 3 for RNN and dense layer, respectively. To improve results of the RNN classifier, we also performed LSTM. The highest classification of 0.969 and 0.983 were attained by RNN and LSTM models, respectively. As the results imply, LSTM has almost same accuracy as the 1-D CNN but with lower number of epochs. Lower number of epochs suggests that LSTM has a lower computational cost. However, a more detailed look at the LSTM graphs, shows some level of instability after the 17th epoch.

### D. The One-One-One deep neural network Model

As discussed above and in section E-V of our methodology, there are significant shortcomings of RNN, CNN and LSTM. Therefore, we proposed the One-One-One Deep Neural Network Architecture to discriminate between people based on their balance abilities. The loss and accuracy graphs, shown in figures 13 (a & b), of train and validation data gave excellent results with significantly high consistency. As shown in the graphs, the validation and train data started to converge at about the 12th epoch and remained totally stable. This level of stability was not observed in any other models despite their high classification performance. Figure 13 (c) shows the receiver operating characteristics graph of the proposed One-One-One Neural Networks classifier performance.

**Figure. 11.**
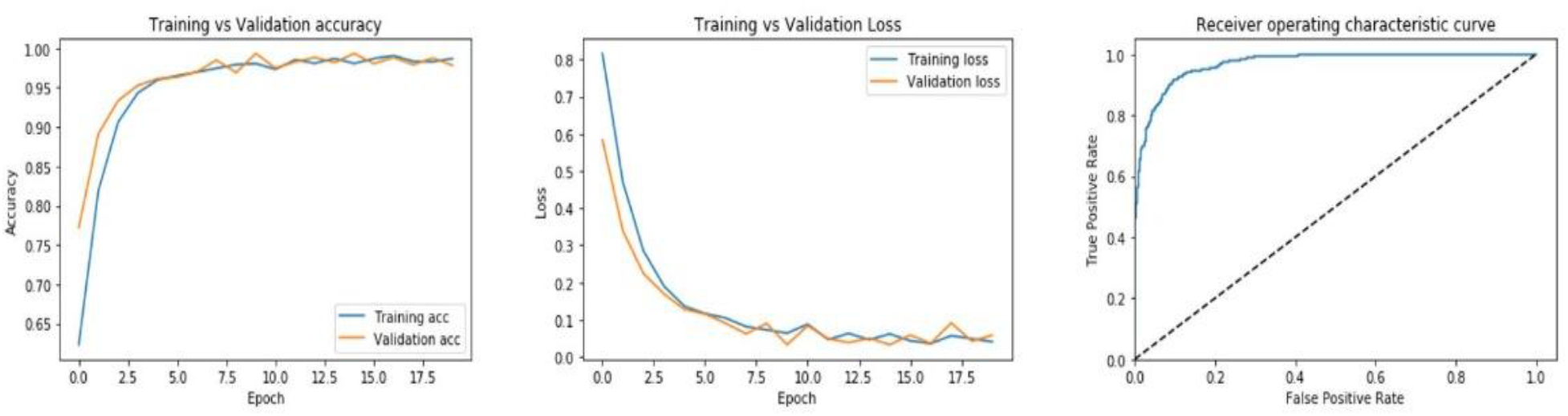
RNN classifier’s Training vs. Validation (a) Accuracy (b) loss (c) Receiver operating characteristics.

**Figure. 12.**
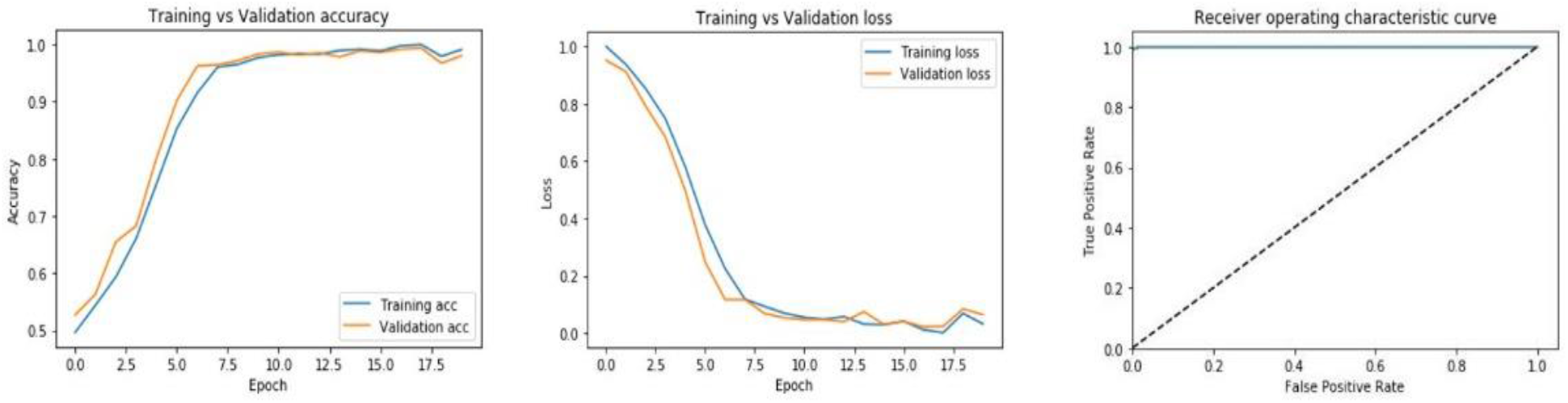
LSTM classifier’s Training vs. Validation (a) Accuracy (b) loss (c) Receiver operating characteristics.

**Figure. 13.**
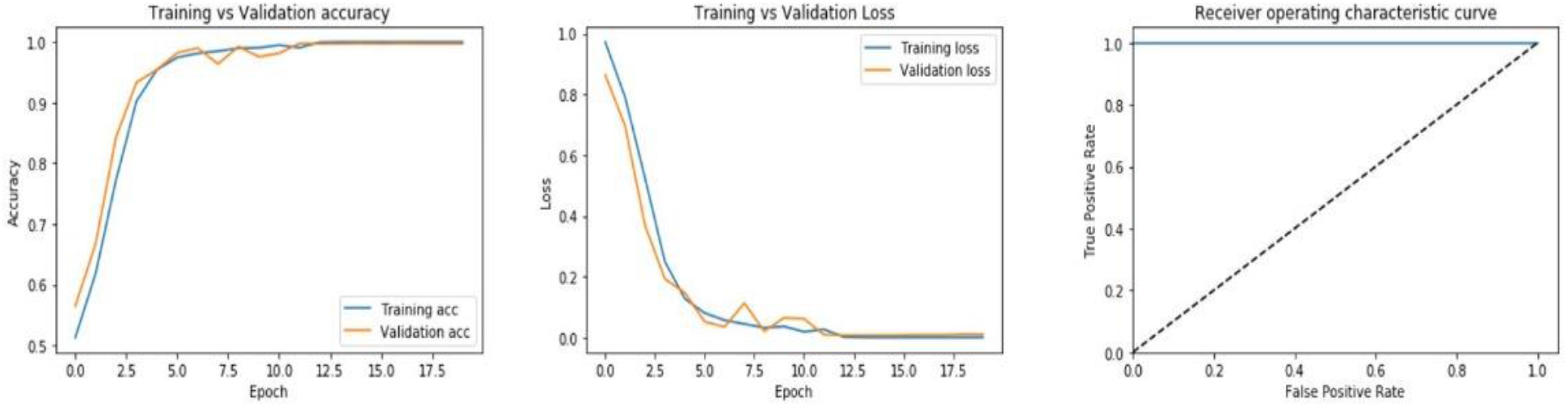
One-One-one Neural Networks classifier’s Training vs. Validation (a) Accuracy (b) loss (c) Receiver operating characteristics.

Table 2 shows the experimental summary using confusion matrix, precision, sensitivity, and accuracy. Based on this table, the experimental result suggests that the proposed model is the most efficient with a precision, sensitivity, and accuracy of 100 %, 100% and 99.9% respectively at the 12^th^ epoch.

As shown on table 2, the proposed One-One-One model provides higher performance and accuracy than more complex systems of One-One-Three and One-One-Two Neural Networks. This suggests that architectural simplicity can be prior to complexity while designing NN models. Thus, the proposed One-One-One model is computationally less expensive for the given input size.

## V. DISCUSSION

Although we got almost the same classification accuracy by all four models, they may not be equally efficient in analyzing the force-plate dataset. As shown in figures (10-11), CNN was time-expensive; it needed at least 50 epochs to experience stability and high system performance of 99.3% accuracy. This may be due to the fact the optimal performance of a CNN architecture is influenced by the number of convolutional layers used. To reduce the influence of multiple layers, we minimized the number of CNN convolutional layers.

RNN gave us an accuracy of 96.9%, as discussed earlier. The low accuracy may be due to the inherent problem of vanishing gradient. We addressed this shortcoming using its variant; LSTM version. LSTM gave a better result, however, at the 17th epoch it shown some degrees of instability; figures 16 and 17. The ineffectiveness and shortcomings of CNN, RNN and LSTM necessitated the design of the of the proposed One-One-One Neural Networks model.

The proposed One-One-One Neural Networks classifier system was designed using only one 1D-convolutional layer on top of one LSTM and one dense layer. Therefore, the proposed approach reduced complexity and improves fitness. For example, using only one 1D CNN layer eliminates the complexity of multiple layers. It also ensures that the model does not remove too many informative features during convolution. In fact, through the following formula we can determine the number of output features by each convolutional layer:

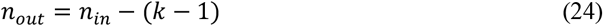

in which *n*_*out*_ is number of output features, *n*_*in*_ is the number of input features and *k* is the kernel size (Dumoulin et al., 2018). Here, we have 21 input features and a 1D-convolutional layer with kernel size:3. Using equation 24, the number of output features *n*_*out*_ is givens as:

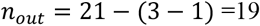

Therefore, only 2 features are removed after data is filtered by 1D-convolutional layer in the proposed One-One-One Neural Networks classifier architecture. Thus, information loss is minimized. Furthermore, the calculated result is consistent with the model summary generated from keras. As shown in table 3, the output of the Conv1d displayed -None, 19, 64.

**Table 3.** Model Summary

| Layer (type) | Output Shape | Param # |
| --- | --- | --- |
| conv1d_2 (Conv1D) | (None, 19, 64) | 256 |
| lstm_2 (LSTM) | (None, 256) | 328704 |
| dense_3 (Dense) | (None, 128) | 32896 |
| dense_4 (Dense) | (None, 3) | 387 |
| Total params: 362,243 |  |  |
| Trainable params: 362,243 |  |  |
| Non-trainable params: 0 |  |  |

The batch_size is not fixed; this is shown as None values in each of the output shapes. The numbers of features output at the Conv1d is shown as 19, this agrees with value we computed using equation 24 above. 64 is the number of filters used.

The param # is the number of parameters or weights that are produced. These parameters are learned during the training of the model. The LSTM and the Dense produced 328 794 and 32 896 parameters, respectively. The large value of the weight produced at the LSTM shows the impact of the 256 filters in learning the temporal nature of the dataset. The “softmax” layer (dense_4) has 3 output shape because our dataset is classified into 3 classes.

The param # of the “softmax” shows 387, this implies that each node in dense_3 (128) is mapped to each of the 3 nodes in the “softmax” layer with an added 3 for the bias: 3 * 128 +3 =387. The total params at the bottom of the table is the sum total of all the parameters learned in the network; 256 + 328 704 + 32 896 + 387 = 362 243. The trainable parameters are the total number of parameters or weights that the networked learned and adjusted their values for the optimization of the model.

As shown in the experimental result, among four models, the proposed One-One-One Neural Networks classifier can be considered as the least computationally expensive model for classification of the human balance dataset in this study. This is because it took only 12 epochs to the reach 99.9% accuracy and maintain stability.

## VI. Conclusion

According to Occam razor principle, simplicity is prior to complexity until proven otherwise (Oladunni & Sharma, 2017). In this work, simple architecture of the proposed One-One-one Neural Networks classifier produced the best result. Thus, eliminated the need for a more complex architecture. Our results also prove that the proposed One-One-One Neural Networks classifier has the capability of extracting the maximum amount of required spatiotemporal information from the force-plate using the randomized sample datasets. The extracted information from the dataset turns out to be necessary and sufficient for training and testing the proposed model with maximum accuracy, sensitivity, and precision without unnecessary architectural complexity. Generalizability of the model was further improved by hyperparameter tuning based on the exhaustive search of the grid for the optimal values of its hyperparameters’ values. Section IV illustrates the effectiveness of this methodology.

The outcome of our experiments (Table 2) shows that we do not have enough evidence to reject our hypothesis. Therefore, we contend that *the proposed one-one-one neural network demonstrated to be the most efficient neural network model in predicting human’s fall-risk using the force-plate time series signal*.

Intuitively, the hybrid of the proposed One-One-One Neural Networks classifier benefits from advantages of both CNN, LSTM and Dense. Therefore, it is logical that the model is effective in analyzing the 1D data with a spatiotemporal structure such as force-plate time series. In fact, CNN does the feature extraction process and prepares data for LSTM which interprets the features across time steps (Brownlee, n.d.; Zhou et al., 2015).

Table 4 compares the performance of our experiment with the state-of-the-art on the same dataset. Balance metrics of 163 subjects were used by different researchers to differentiate people based on their gender, age, risk of fall, etc. We focused on a non-binary classification and discriminated people based on their balance abilities not only on their gender or age as done in previous studies.

**Table 4.** Comparison with the state-of-the-art

| Study | Year | Best Result (Acc%) |
| --- | --- | --- |
| RF: with six (temporal, spatial, spectral) features used in random selection (Giovanini et al., 2018) | 2018 | 64.9 |
| MLP; used 18 time and frequency domain features by SAFE feature extraction method (Reilly, 2019) | 2019 | 80 |
| SVM; used force signals as features (Cetin & Bilgin, 2019) | 2019 | 81.67 |
| **CNN (ours); used all force, moment, and CoP features** | 2020 | **99.3** |
| **RNN (ours); used all force, moment, and CoP measures** |  | **96.9** |
| **LSTM (ours); used all force, moment, and CoP measures** |  | **98.3** |
| **One-One-Three NN (ours); used all force, moment, and CoP measures** |  | **99.5** |
| **One-One-Two NN (ours); used all force, moment, and CoP measures** |  | **99.7** |
| **One-One-One NN (Proposed); used all force, moment, and CoP measures** |  | **99.9** |

Based on the outcome of our experiment, we argue that the proposed model is reliable and efficient in predicting risk of fall in human subjects of different ages based on their fall concern and balance abilities. In other words, this model can detect balance impairment in different range of people with age, gender, health status, fall history, illness, medication use, impairment background and other specifications. The dataset for this work was retrieved from (dos Santos & Duarte, 2016; Santos & Duarte, 2016) and contains the balance characteristics of many human subjects recorded by a force-plate.

This study shows the effectiveness and performance of deep neural networks in building an accurate predictive model. Its effectiveness without the feature extraction stage of the traditional machine learning is evident as compared with our previous experiments (Savadkoohi et al., 2020). The promising result of the present study is a motivation in exploring the architecture of the proposed One-One-One Deep Neural Networks in discerning patterns and discovering knowledge in other scientific problems. However, experimental result may not be the same for all datasets. This is a limitation of this work because architectural simplicity and minimum complexity approach may not be adequate for all problems.

Major contributions of this work are as follows.

a. Table 4 shows that the worst classification accuracy of our work, 96.9% using RNN, is far above and beyond the highest accuracy of 80 and 81.67% achieved by other researchers in 2019 (Cetin & Bilgin, 2019; Reilly, 2019). To the best of our knowledge, we do not find any other study on this dataset with higher classification results. Therefore, we consider this work as the new baseline.
b. 4 deep neural network models were designed, developed, and evaluated to predict human’s balance impairment using the force-plate time series signal. With an accuracy of 99.9% at the 12^th^ epoch, our experiment shows that the proposed One-One-One Neural Networks has computational cost advantage over other models.
c. Optimization of the proposed One-One-One Neural Networks classifier architecture was based on a combination of: i) random sampling for data selection, ii) architectural simplicity and minimum complexity approach, and iii) Exhaustive search technique of hyperparameters’ values using the grid searching methodology.
d. Classification was based on the Short Falls Efficacy Scale International test (FES) for the identification of individuals’ concern (fear) of a fall. Subjects were classified as low, moderate, and high concern groups due to their FES test results which is a standard test to measure the fall risk.

The implications of the study are as follows:

1. Balance impairment in human is predictable using deep neural networks. Our experiment employed this state-of-the-art learning algorithms to classify force-plate balance time-series signal to predict human’s balance impairment. Classification was based on low, moderate, and high risk of a fall.
2. Combining one layer of 1D CNN with one layer of LSTM and a Dense layer produces the most efficient neural network model in predicting human’s balance impairment using the force-plate time series signal. The proposed One-One-One Neural Networks classifier model provides a considerable increase in performance compared to other algorithms. Our experimental results show precision, sensitivity, and accuracy of 100 %, 100% and 99.9% respectively at the 12^th^ epoch.
3. Ultimately, using faster computers with higher CPU and more powerful processors, can lead to achieving more insightful perspective and gain a new knowledge regarding the human balance underlying patterns and its characteristics.
4. Our experiment suggests that architectural simplicity and minimum complexity approach is critical in building efficient deep neural networks. The proposed One-One-One Neural Networks classifier demonstrated the capability of extracting maximum spatiotemporal information from the randomized sample datasets. The extracted information turned out to be necessary and sufficient for training and testing the proposed model. The outcome of our experiment showed the effectiveness of the strategy.

In the future, we will consider the following:

1. A multiclass identification of individuals’ concern (fear) of a fall into very low, low, moderate, high, and very high.
2. Dimensionality reduction of the explanatory variables to improve computational efficiency.

## Data Availability

Data used in this study is a publicly available dataset. It is accessible through both PhysioNet (DOI: 10.13026/ C2WW2W) and Figshare (DOI: 10.6084/ m9. figshare. 3394432) websites.

https://physionet.org/content/hbedb/1.0.0/

https://figshare.com/articles/dataset/A_public_data_set_of_quantitative_and_qualitative_evaluations_of_human_balance/3394432

## VIII. Acknowledgment

We would like to acknowledge the University of the District of Columbia. Further, we would like to acknowledge the Department on Aging and Community Living (DACL) for their sponsorship. We also like to acknowledge the following federal funding sources: National Science Foundation (NSF) Award #’s 2032345, 1654474, and1700219.

